# Impact of the Coronavirus Disease (COVID-19) on the Mental Health and Physical Activity of Pharmacy Students at the University of Zambia: A Cross-Sectional Study

**DOI:** 10.1101/2021.01.11.21249547

**Authors:** Steward Mudenda, Moses Mukosha, Chiluba Mwila, Zikria Saleem, Aubrey Chichoni Kalungia, Derick Munkombwe, Victor Daka, Bwalya Angel Witika, Martin Kampamba, Misheck Chileshe, Christabel Hikaambo, Maisa Kasanga, Webrod Mufwambi, Ruth Lindizyani Mfune, Scott Kaba Matafwali, Angela Gono Bwalya, David Chimbizgani Banda, Akash Gupta, Maureen Nkandu Phiri, Eustarckio Kazonga

## Abstract

**Background:** The novel coronavirus disease (COVID-19) is a serious global health problem that has negatively impacted the mental health of students.

**Methods:** We conducted an online descriptive cross-sectional study among 273 undergraduate pharmacy students at the University of Zambia. A partial proportional odds regression model was used to determine the predictors of anxiety. All statistical tests were set at 95% confidence level (p<0.05).

**Results:** A response rate of 70% was obtained with the majority of the students being female 51.6%. Of the 273 respondents, 23.8% did not experience anxiety, 34.4% experienced mild anxiety, 24.9% experienced moderate anxiety while 16.9% experienced severe anxiety about COVID-19. It was also found that 61.2% of students reported that their attention to mental health increased during the COVID-19 pandemic whereas 44.3% reported an increased resting time with a significant reduction in relaxation 51.3% and physical activity 45.4% time. Factors that affected mental health included; reduced family care (OR: 2.27; 95% CI: 1.09-4.74), not changing attention to mental health (OR: 0.33; 95% CI: 0.18-0.62), being in the final year of study (OR: 0.33; 95% CI: 0.13-0.84), reduced time of resting (OR: 2.10; 95% CI: 1.26-3.50) and feeling helpless (OR: 0.42; 95% CI:0.23-0.75).

**Conclusion:** COVID-19 negatively impacted the mental health and physical activity of pharmacy students at the University of Zambia. This can have negative health and academic outcomes for students going forward. Higher learning institutions and key stakeholders should implement measures to aid students to recover from the impact of COVID-19 on their mental health and physical activity.

## Introduction

The novel coronavirus disease (COVID-19) originated from Wuhan city, Hubei province of China [1], with 27 cases of pneumonia of unknown cause identified on 31^st^ December 2019 [2,3]. The causative agent of this infection was later named Severe Acute Respiratory Syndrome Coronavirus 2 (SARS-CoV-2) [2,4,5]. This led to the naming of the disease as COVID-19 [6]. The main clinical presentations of patients suffering from COVID-19 at that time included dry cough, sore throat, chest pain, dyspnoea, headache, fever, nausea, vomiting, diarrhoea, bilateral lung infiltration on imaging, and abnormal radiological findings [7–10]. Current findings have reported clinical features such as myalgias, fatigue, abdominal pain, tachycardia, tachypnea, and hypotension [11]. COVID-19 was declared a global pandemic by the World Health Organization (WHO) on 11^th^ March 2020 due, in part, to its rapid spread across the globe and relatively high mortality rate among specific populations [6,12–14]. As a result, many countries instituted lockdowns to prevent further spreading of COVID-19 [15,16].

There is strong evidence suggesting that COVID-19 has had adverse effects on the mental wellbeing of individuals worldwide [17–21]. Some of the mental health problems observed were illness anxiety disorder (IAD or formerly hypochondriasis), helplessness and horror, and the fear of COVID-19 associated stigmatization [22–25]. Besides, some populations have reportedly experienced psychological impacts including but not limited to; depression, anxiety, stress, and mood swings due to the pandemic [18,22,26–29].

Some studies have shown that there is a positive relationship between physical activity and mental health [30]. Physical activity plays a significant role in reducing the symptoms of mental health illnesses [31]. Lockdowns and restrictions to movement during the COVID-19 pandemic have affected the time that people spend resting, relaxing, and exercising [32,33]. Many people experienced changes in their usual day-to-day routines, including physical activity, and that may have contributed to mental health disorders in most populations [34].

As of 1^st^ December 2020, the number of COVID-19 cases in Zambia was approximated at 17,647 with 357 deaths [35]. The rapid spread of COVID-19 and aforementioned lockdowns led to the closure of schools, colleges, and universities worldwide [36,37]. This was done to prevent further transmission and spread of the virus [12,38]. Unfortunately, the closure of learning institutions has been reported to negatively affect the mental health and academic life of students [39,40]. Many youths, the population of which the majority of the students comprises, have experienced mental health problems due to COVID-19 [39]. Recent studies have reported that students have experienced symptoms of anxiety, depression, stress, panic, and mood swings due to COVID-19 and its negative effects on the education system [41–43]. This may in turn affect the academic performance of the students compared to their performance before the COVID-19 era [44,45]. COVID-19 has disrupted many educational and extracurricular activities for students [43,46,47]. However, most of the impacts of COVID-19 on academic performance will be known during the post-COVID-19 era.

A literature search did not yield previous studies on the impact of COVID-19 on the mental health and physical activity of students and other populations in Zambia. Thus, as an initial step, we assessed the impact of COVID-19 on the mental health and physical activity of undergraduate pharmacy students at the University of Zambia (UNZA) and reported them *vide infra*. This was against a background that training of pharmacists in Zambia has over the years largely been utilising traditional face-to-face teaching and learning, including experiential learning in pre-clinical and clinical subject areas [48]. The restrictions and health risks imposed by COVID-19 presented unique challenges not only for the facilitation of learning in schools and universities but also to psychosocial wellbeing of the learners. Therefore, our study aimed to assess the impact of COVID-19 on the mental health and physical activity of undergraduate pharmacy students at the University of Zambia.

## Methods

### Study Design, Participants and Sampling

This was a descriptive cross-sectional study involving the undergraduate pharmacy students at UNZA – the largest and leading public university in Zambia. Thus, the results of this study gave an insight into the mental health and physical activity impact of COVID-19 on pharmacy students. The study included all the enrolled cohorts of undergraduate pharmacy students at UNZA, School of Health Sciences, that provided consent to take part in the study during the academic year 2019 to 2020. UNZA is currently the major producer of Bachelor of Pharmacy graduates in Zambia [48]. Undergraduate pharmacy students without access to the online questionnaire were excluded from the study.

The study population was 410 pharmacy students, determined as follows: a sum of 90 second year, 140 third year, 126 fourth year, and 54 fifth-year pharmacy students, cumulatively. The sample size was determined using Yamane’s formula; 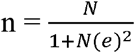 [49]. A margin of error of 5% and a population of 410 were used. A 10% loss or non-response was anticipated. However, 288 students responded representing a response rate of 70%. From the 288 pharmacy students, a total of 273 students managed to complete the questionnaire. 15 students did not complete responding to the questionnaire.

### Data Collection Tool

Data were collected using an online self-administered questionnaire that was adapted from similar studies [39,41,50]. Firstly, the questionnaire was circulated to experts from the University of Zambia to allow for content and face validation. The modified questionnaire was pre-tested among 10 undergraduate pharmacy students at UNZA, who, later were excluded from the main study. Student’s anxiety was measured using the generalised anxiety disorder 7-item (GAD-7) scale developed by Spitzer *et al*., [51]. The GAD-7 scale is used to categorize anxiety into four classes, namely; no anxiety, mild, moderate, and severe anxiety. Scoring of GAD-7 anxiety levels was done from 0 to 21 and categorised as follows; 1-4 no anxiety, 5-9 mild anxiety, 10-14 moderate anxiety, and 15-21 severe anxiety. The questionnaire was used to collect data on socio-demographic characteristics and mental health and physical activity impacts of COVID-19 on undergraduate pharmacy students from 1^st^ August to 30^th^ September 2020.

### Study outcome

Mental health: we evaluated mental health using the four levels of anxiety disorders from the GAD-7 anxiety levels as follows

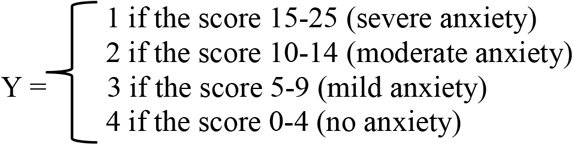

### Data Analysis

Data were analysed using Stata/IC version 16 (Stata Corp., College Station, Texas, USA). The significance level was set at alpha 0.05 and 95% confidence intervals.

For descriptive statistics, the median (interquartile range [IQR]) for continuous values (i.e age) was calculated after testing for the assumption of normality using Shapiro-Wilk W-test and graphically qq-plots. For the comparison of baseline characteristics across levels of anxiety, we used the Kruskal Wallis test and Pearson chi-square test or Fisher’s exact test were appropriate

In order to choose a model to identify independent predictors of anxiety as a response with four ordered categories. firstly the confounders to be included in the model were found by carrying out backward elimination using the socio-demographic variables as candidates with a liberal p-value (0.20) for exclusion, in order to ensure that all possible confounders were included. After carrying out the backward elimination, forward selection (with the same liberal P-value of 0.20 for inclusion) was carried out to see whether any other possible confounders would be identified. In this case, both backward elimination and forward selection identified the same model.

To get some idea of the effect of academic, social and family support factors, Proportional Odds Models (POM) were then fitted for each of the factors separately, adjusting for the socio-demographic variables that had been identified. Thereafter backward elimination was carried out with a stricter P-value for the exclusion of p=0.10 with age fixed in the model, irrespective of whether it was still statistically significant or not. The model was then refitted and tested for proportional odds assumption using the brant test. The model was found to be violating the proportional odds assumption and therefore a partial Proportional Odds Model (PPOM) was fitted. Each model’s fit was evaluated using the deviance test as well as the log-likelihood and the pseudo R2.

### Ethical Considerations

This study was approved by the University of Zambia Health Sciences Research Ethics Committee (UNZAHSREC). **Protocol ID**: 2020310174. **IORG no: 0009227 IRB no: 00011000 FWA no: 00026270**. After IRB ethical approval, regulatory approval was obtained from the National Health Research Authority (NHRA).

## Results

### Socio-demographic characteristics of study participants

The baseline characteristics of respondents by levels of anxiety are shown in table 1. A total of 273 respondents with a median age of 24 years (IQR, 22-27) were included in the analysis. From the total, about 65 (23.8%) had no anxiety, 94 (34.4%) had mild anxiety, 68 (24.9%) hard moderate anxiety and 46 (16.9%) had severe anxiety. There were more females than males 141 (51.6%). The majority were single 225 (82.4%), were in the third year of study 113 (41.4%), lived in urban areas 156 (57.1%), lived with parents 178 (65.2%) and were unemployed 196 (71.8%). From the univariate analysis, there was evidence of a difference in residence (P=0.005), marital status (p=0.024) and whether or not a respondent was living with their parents (P=0.022), across all the levels of anxiety.

**Table 1.** Socio-demographic characteristics of participants by levels of anxiety.

| Characteristic | Levels of anxiety, N=273 |  |  |  | P-value |
| --- | --- | --- | --- | --- | --- |
|  | No anxiety<br>n=65 (23.8%) | Mild anxiety<br>n=94 (34.4%) | Moderate anxiety<br>n=68 (24.9%) | Severe anxiety<br>n=46 (16.9%) |  |
| **Age median (IQR)** | 24 (23-28) | 23.5 (22-25) | 23 (22-26.5) | 24 (22-28) | 0.195<sup>a</sup> |
| **Sex, n (%)** |  |  |  |  |  |
| *male* | 31 (23.5) | 49 (37.1) | 30 (22.7) | 22 (16.7) | 0.791<sup>b</sup> |
| *female* | 34 (24.1) | 45 (31.9) | 38 (26.9) | 24 (17) |  |
| **Marital status, n (%)** |  |  |  |  |  |
| *single* | 48 (21.3) | 85 (37.8) | 57 (25.3) | 35 (15.6) | 0.024<sup>c</sup> |
| *married* | 17 (36.2) | 9 (19.2) | 11 (23.4) | 10 (21.3) |  |
| **Year of study, n (%)** |  |  |  |  |  |
| *Second* | 4 (13.3) | 8 (26.7) | 10 (33.3) | 8 (26.7) | 0.102<sup>c</sup> |
| *Third* | 26 (23) | 36 (31.9) | 32 (28.3) | 19 (16.8) |  |
| *Fourth* | 17 (21) | 30 (37.5) | 17 (21.2) | 16 (20) |  |
| *Fifth* | 18 (36) | 20 (40) | 9 (18) | 3 (6) |  |
| **Employed , n (%)** |  |  |  |  |  |
| *Yes* | 23 (29.9) | 21 (27.3) | 16 (20.8) | 17 (22.1) | 0.128<sup>b</sup> |
| *No* | 42 (21.4) | 73 (37.2) | 52 (26.5) | 29 (14.8) |  |
| **Residence, n (%)** |  |  |  |  |  |
| *Urban* | 44 (28.2) | 59 (37.8) | 30 (19.2) | 23 (14.7) | 0.005<sup>b</sup> |
| *Rural-urban* | 18 (27.3) | 16 (24.2) | 18 (27.3) | 14 (21.2) |  |
| *Rural* | 3 (5.9) | 19 (37.3) | 20 (39.2) | 9 (17.7) |  |
| **Living with parents, n (%)** |  |  |  |  |  |
| *Yes* | 34 (19.1) | 70 (39.3) | 47 (26.4) | 27 (15.2) | 0.022<sup>b</sup> |
| *No* | 31 (32.6) | 24 (25.3) | 21 (22.1) | 19 (20) |  |

Key: <sup>a</sup>=Kruskal Wallis test, <sup>b</sup>=Pearson chi-square test, <sup>c</sup>=Fisher's exact test, IQR-interquartile range

### Academic life, social networks and family support by levels of anxiety

Table 2 shows the academic life, social networks and family support by levels of anxiety. We noted that there were more respondents with no steady income 187 (68.5%), with reduced social support from family 118 (43.5%), with reduced social support from friends 182 (66.7%), who were feeling helpless 189 (69.2%), had increased resting time 120 (44%), with reduced relaxation time 140 (51.3%), with reduced academic performance 221 (81%) and with reduced physical activity 124 (45.4%). Across all levels of anxiety, there was a statistically significant difference in steady income (p=0.014), social support from family (p=<0.0001), social support from friends (p=0.026), helpless (p=<0.0001), resting time (p=0.001), relaxing time (p=0.002), academic performance (p<0.0001) and exercise (p=0.017).

**Table 2.** Academic life, social networks and family support of participants by levels of anxiety.

| Characteristic | Levels of anxiety, N=273 |  |  |  | P-value |
| --- | --- | --- | --- | --- | --- |
|  | No anxiety<br>n=65 (23.8%) | Mild anxiety<br>n=94 (34.4%) | Moderate anxiety<br>n=68 (24.9%) | Severe anxiety<br>n=46 (16.9%) |  |
| **Feeling helpless, n (%)** |  |  |  |  |  |
| Yes | 29 (15.3) | 64 (33.9) | 58 (30.7) | 38 (20) | <0.0001<sup>b</sup> |
| No | 36 (42.9) | 30 (35.7) | 10 (11.9) | 8 (9.5) |  |
| **Time resting, n (%)** |  |  |  |  |  |
| Has increased | 34 (28.1) | 44 (36.6) | 25 (20.7) | 18 (15) | 0.001<sup>b</sup> |
| Has reduced | 11 (10.5) | 37 (35.2) | 34 (32.4) | 23 (21.9) |  |
| Remain same | 20 (42.6) | 13 (27.7) | 9 (19.1) | 5 (10.6) |  |
| **Time relaxing, n (%)** |  |  |  |  |  |
| Has increased | 25 (26.6) | 38 (40.4) | 18 (19.2) | 13 (13.8) | 0.002<sup>b</sup> |
| Has reduced | 22 (15.7) | 47 (33.6) | 44 (31.4) | 27 (19.3) |  |
| Remain same | 18 (46.2) | 9 (23.1) | 6 (15.4) | 6 (15.4) |  |
| **Time exercise, n (%)** |  |  |  |  |  |
| Has increased | 16 (22.9) | 32 (45.7) | 11 (15.7) | 11 (15.7) | 0.017<sup>b</sup> |
| Has reduced | 27 (21.8) | 35 (28.2) | 33 (26.6) | 29 (23.4) |  |
| Remain same | 22 (27.9) | 27 (34.2) | 24 (30.4) | 6 (7.6) |  |
| **Academic performance, n (%)** |  |  |  |  |  |
| Has increased | 11 (55) | 3 (15) | 2 (10) | 4 (20) | <0.0001<sup>c</sup> |
| Has reduced | 38 (17.2) | 83 (37.6) | 62 (28.1) | 38 (17.2) |  |
| Remain same | 16 (50) | 8 (25) | 4 (12.5) | 4 (12.5) |  |
| **Family Support, n (%)** |  |  |  |  |  |
| Has increased | 18 (20.9) | 28 (32.6) | 26 (30.2) | 14 (16.3) | <0.0001<sup>b</sup> |
| Has reduced | 20 (17) | 43 (36.4) | 28 (23.7) | 27 (22.9) |  |
| Remain same | 26 (38.8) | 23 (34.3) | 13 (19.4) | 5 (7.5) |  |
| **Friends support, n (%)** |  |  |  |  |  |
| Has increased | 7 (20) | 12 (34.3) | 11 (31.4) | 5 (14.3) | 0.026<sup>b</sup> |
| Has reduced | 36 (19.8) | 61 (33.5) | 48 (26.4) | 37 (20.3) |  |
| Remain same | 22 (39.3) | 21 (37.5) | 9 (16.1) | 4 (7.1) |  |
| **Prefer E-learning, n(%)** |  |  |  |  |  |
| Yes | 21 (36.2) | 15 (25.9) | 12 (20.7) | 10 (19.2) | 0.077<sup>b</sup> |
| No | 44 (20.5) | 79 (36.7) | 56 (26.1) | 36 (16.7) |  |
| **Steady income, n (%)** |  |  |  |  |  |
| Yes | 30 (34.9) | 27 (31.4) | 14 (16.3) | 15 (17.4) | 0.014<sup>b</sup> |
| No | 35 (18.7) | 67 (35.8) | 54 (28.9) | 31 (16.6) |  |

Key: <sup>a</sup>=Kruskal Wallis test, <sup>b</sup>=Pearson chi-square test, <sup>c</sup>=Fisher's exact test, IQR-interquartile range

### Factors affecting the mental health of undergraduate pharmacy students

Table 3 shows the results of the Partial Proportional Odds regression Model (PPOM). The log-likelihood ratio chi-square test LRX^2^(19)=95.77, p<0.0001 for this model indicates that the full model shows a better fit than the null model with no independent variables in predicting the levels of anxiety. The Wald test of parallel lines assumption for the final model: Chi-square test = 29.27, df=32, p=0.606 indicates that the final model does not violet the proportional odds assumption. Since academic performance violates the PO assumption its effect was allowed to vary across the three binary models.

**Table 3:**
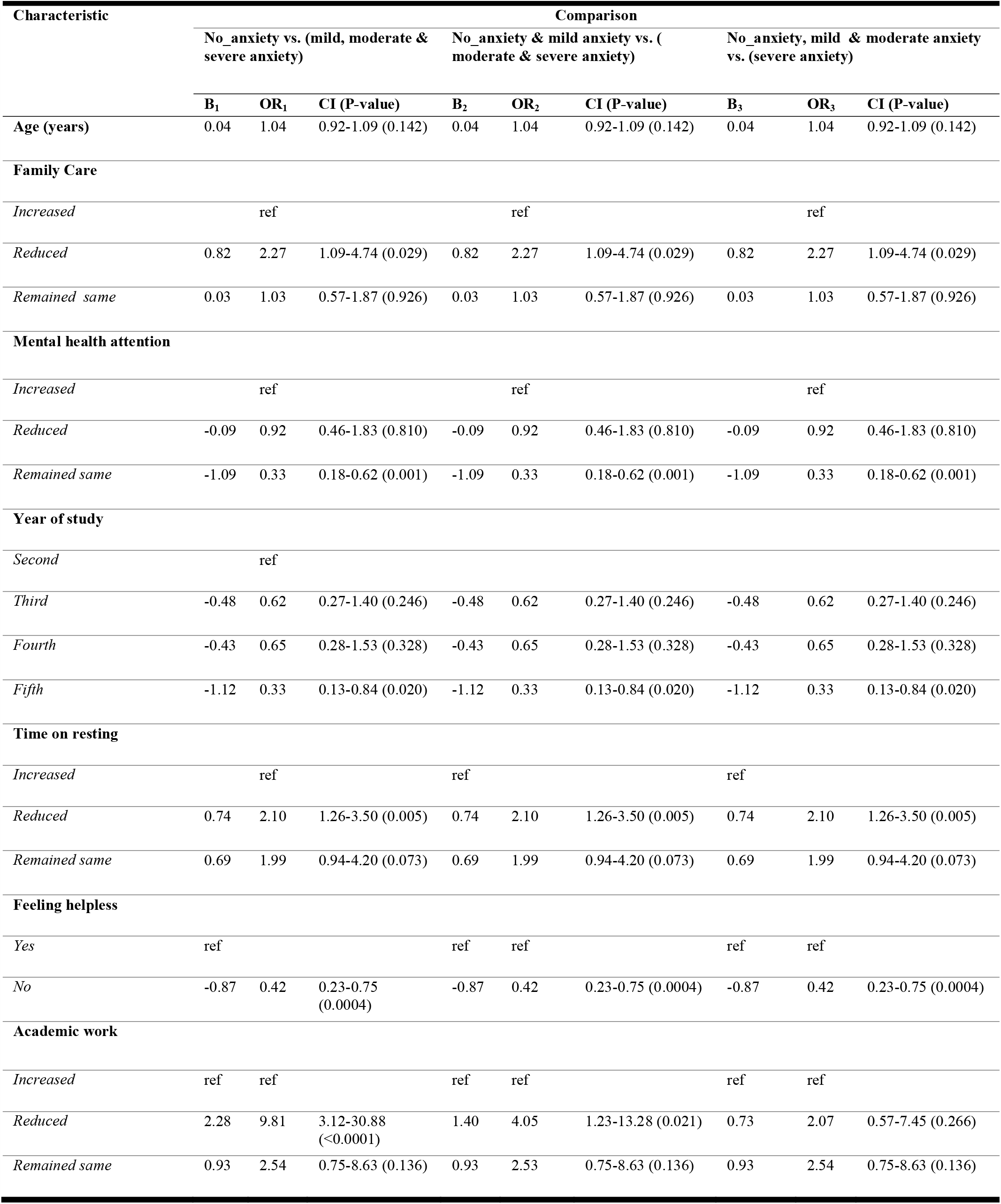
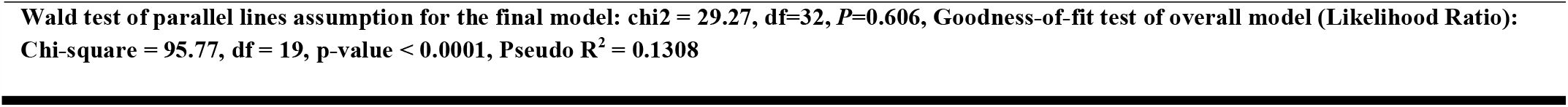
Factors affecting the mental health of undergraduate pharmacy students.

This model indicated that the factors associated with levels of anxiety among the respondents were reduced care from family, not changing attention to one’s mental health, being in the final year of study, reduced time of resting and feeling helpless.

Compared to increased family care, those who reported receiving reduced family care (aOR: 2.27, 95% CI: 1.09-4.74, p=0.029) and reduced resting time (aOR: 2.10, 95% CI:1.26-3.50, p=0.005) were 27% and 10% more likely to be in higher levels of anxiety (i.e severe anxiety) respectively adjusting for all covariates. The respondents who were in their final year of studies compared to the second year (aOR:0.33, 95% CI: 0.13-0.84, p=0.020) and those not feeling helpless (aOR: 0.42, 95% CI: 0.23-0.75) were 67% and 58% less likely to be in higher levels of anxiety respectively when holding other covariates constant. Meanwhile, a unit increase in age was associated with increased odds of being beyond a particular category of anxiety (higher category) by a factor of 1.04 though we could not rule out random chance finding (p=0.142). Overall academic performance (i.e reduced compared to increased) was associated with 81% and 40% increase in the likelihood of a respondent being in higher levels of anxiety for the first (95% CI: 3.12-30.88, p<0.0001) and second (95% CI: 1.23-13.28, p=0.021) binary models. On the contrary, we noted a 27% reduced likelihood of being in higher levels of anxiety across the third binary model (95% CI: 0.57-7.45) but this was not statistically significant (p=0.266).

## Discussion

We assessed the impact of COVID-19 pandemic on the mental health and physical activity of undergraduate pharmacy students at the University of Zambia. We found that the factors associated with levels of anxiety among the students were reduced care from family, not changing attention to one’s mental health, being in the final year of study, reduced time of resting and feeling helpless. Currently, COVID-19 has caused many negative impacts on the education sector and other sectors across the globe.

### Mental health

Our study established that COVID-19 has affected the mental health of pharmacy students and reduced their time to do physical activity. Though this was expected, it is the magnitude of the impact on mental health that is noteworthy. Our study found that 76.2% of the students experienced the anxiety of which the majority (34.4%) had experienced mild anxiety. Our survey further found that many students felt helpless (69.2%) as a result of the COVID-19 pandemic spreading in Zambia. Previous studies have also reported that COVID-19 affects the mental health of students. A study in China reported that most of the students (21.3%) experienced mild anxiety [41]. Another study in China reported overall anxiety of 26.0% with the majority of the students (23.2%) experiencing mild anxiety [52]. The study further reported that students had experienced depression due to the impact of COVID-19 [52]. Xiao *et al*.reported that anxiety prevalence among medical students was 17.1% and depression was 25.3% [53]. In Bangladesh, Khan *et al*.(2020) reported that 33.3% of the students experienced anxiety [54]. Further, other mental health disorders such as depression, stress, and event-specific distress were reported. Similarly, in Pakistan, Waseem *et al*. (2020) reported that 53.0% of undergraduate medical students experienced stress as a result of COVID-19 [42]. Our survey and other similar studies reported that students experienced mental health problems associated with COVID-19. However, the prevalence of mental health problems reported in our study is higher than the ones reported in similar studies. This difference could be because our setting is resource-constrained. Also, the prevalence of anxiety in our study may have been higher since the pharmacy students experienced online learning for the first time.

The mental health problems reported in our study were mainly associated with reduced care from family members, not changing attention to one’s mental health, being in the final year of study, reduced time of resting and feeling helpless. Similar studies reported that mental health problems could have been affected by factors such as fear of contracting the virus [55], social distancing [53], economic impacts of COVID-19 [56], fear of delayed academic progression [41], fear of poor academic performance [45], and impact of COVID-19 on family and social networks [39].

Mental health problems associated with COVID-19 have been reported in different populations including healthcare workers [57,58] and the general population [59]. Therefore, this indicates that COVID-19 has impacted the mental health of different populations of people.

### Physical activity and mental health

In this survey, the majority of the students’ attention to mental health (61.2%) and time to rest (44.3%) has increased as a result of COVID-19. However, most of the students reported that their time to relax (51.3%) and exercise (45.4%) had reduced. It was found that the students spent more time resting, but very little time relaxing and exercising. Physical activity has been linked to the improved mental health of human beings [60–63]. Additionally, physical activity reduces mental health problems such as anxiety, depression, social withdrawal, low self-esteem, and mood swings [64,65]. Even minimal physical activities while staying at home can be performed so that individuals remain active during the pandemic [66,67]. However, due to COVID-19, preventive measures such as social distancing and restrictions in movements meant that students spent their time at home and had no access to physical exercising facilities like a gymnasium. This caused their resting time to increase while the exercising time reduced and thus may have resulted in anxiety. Similarly, other studies reported that there has been a significant reduction in physical activity as a result of quarantine [61,66,68]. This reduction in physical activities is one of the contributing factors to the reported mental health problems due to COVID-19. There is evidence that indicates that reduced physical activity increases the risks of mental health problems among students [64]. Since many schools, colleges, and Universities are now open in Zambia, students must practice simple exercises while observing the preventive measures of COVID-19.

### Implications on educational policy and practice

The importance of good mental health and wellbeing in cognitive development cannot be understated. With existing evidence showing that the majority of undergraduate pharmacy bstudents at UNZA predominantly utilise a strategic approach to learning - an attribute characterised by a focus on achieving and fear of failure [48], it was a cause for concern that the negative impacts of COVID-19 on the mental health of students could further affect their cognitive development, approaches to learning, and academic outcomes. We, therefore, argue that that educational policy and practice amidst the COVID-19 pandemic and beyond should consider adopting an instructional design that influences and enhances mental resilience in addition to the attainment of meaningful learning outcomes, critical thinking and effective study skills among pharmacy students [48]. We further contend that in addition to modifying the above facets of the learning environment at the university, extra consideration should be given to provide psychosocial support to students negatively affected by the COVID-19 pandemic.

### Strengths and limitations of the Study

This is the first study to assess the impact of COVID-19 on the mental health and physical activity among Zambian students and it has highlighted the need to pay attention to the mental health of students not only during COVID-19 pandemic but even beyond. Further, the study did not only indicate the prevalence of anxiety among pharmacy students due to COVID-19, but it also explored the factors that independently predict the anxiety levels among these students.

Since we conducted the study among pharmacy students, the results cannot be generalised to other students doing different programmes. The online nature of the study meant that certain students had no access to the questionnaire and were, therefore, not part of the study. The study findings may not represent the future academic outcomes of undergraduate pharmacy students.

## Conclusion

This study found that the COVID-19 negatively impacted the mental health and physical activity of undergraduate pharmacy students at the University of Zambia. Our study established that students experiencing anxiety associated with COVID-19 risk were more likely to have poor academic outcomes and progression. Institutions of higher learning and other stakeholders must, therefore, put in place measures to mitigate the negative impact of COVID-19 on the mental health and academic life of students. We will continue to monitor this and other post-COVID-19 effects on educational processes and outcomes in future.

## Data Availability

All data will be availed to any readers who may need it

## What is known about this topic

- Literature search indicates that students from various countries and pursuing different programmes have been experiencing anxiety associated with COVID-19.
- COVID-19 has impacted the education sector and other sectors negatively.

## What this study adds

- This study adds valuable information to existing knowledge on the possible impact of COVID-19 on the mental health and physical activity of students.
- The study adds basic information on COVID-19 vs mental health and COVID-19 vs physical activity.
- This study also adds information on the imapact of COVD-19 on academic progression and performance of students, and thus calls for schools and key stakeholders to provide measures that will help students recover from the negative impact of COVID-19.

## Acknowledgements

The authors are grateful to all the pharmacy students who took their time to participate in this study. We are also grateful to the University of Zambia Library for providing access to the majority of the articles that were used in this study. We thank the following individuals for helping in editing the manuscript; Dr. Jimmy Hangoma, Mr. Moses Ngazimbi, Mr. Frank Mudenda, Ms. Dainess Hang’andu, Mr. Paul Masebe, and Mr. Edgar John Sintema.

## Authors’ contributions

SM conceptulaized the study. Data collection was done by SM, MNP, and CM. Data analysis was conducted by SM, MM, MC, VD, and EK. All authors (SM, MM, CM, XS, ACK, DM, BAW, VD, RLM, MC, MK, AGB, DCB, MK, CH, SKM, AG, WM, MNP, and EK) contributed to data interpretation and writing of the initial manuscript. All authors reviewed the intellectual content of the manuscript. All authors read and reviewed the materials cited in this manuscript. Project administration and supervision was done by SM. All authors read and approved the final version of the manuscript.

